# Climatic influence on the magnitude of COVID-19 outbreak: a stochastic model-based global analysis

**DOI:** 10.1101/2020.06.02.20120501

**Authors:** Malay Pramanik, Koushik Chowdhury, Md Juel Rana, Praffulit Bisht, Raghunath Pal, Sylvia Szabo, Indrajit Pal, Bhagirath Behera, Qiuhua Liang, Sabu S. Padmadas, Parmeshwar Udmale

## Abstract

This study examines the association between community transmission of COVID-19 cases and climatic predictors, considering travel information and annual parasite index across the three climatic zones, i.e., tropical, subtropical, and temperate. A Boosted Regression Tree model has been employed to understand the association between the COVID-19 cases. The results show that average temperature and average relative humidity are the major contributors in explaining the differentials of COVID-19 transmission in temperate and subtropical regions whereas the mean diurnal temperature range and temperature seasonality are the most significant determinants in tropical regions. The average temperature is the most influential factor affecting the number of COVID-19 cases in France, Turkey, the US, the UK, and Germany, and the cases decrease sharply above 10°C. Among the tropical countries, India found to be most affected by mean diurnal temperature, and Brazil fazed by temperature seasonality. Most of the temperate countries like France, USA, Turkey, UK, and Germany with an average temperature between 5–12°C had high number of COVID-19 cases. The findings are expected to add to the ongoing debates on the influence of climatic factors influencing the number of COVID-19 cases and could help researchers and policymakers to make appropriate decisions for preventing the spread.

**Highlights:**

1. Analyzed influence of climatic & bioclimatic factors on the spread of COVID-19
2. First to analyze COVID-19 cases in 228 cities globally across three climatic zones
3. Temperature & humidity influenced COVID-19 cases in temperate & sub-tropics
4. Mean diurnal temperature & temperature seasonality had effects in tropics
5. Low temperature elicits COVID-19 cases in France, Turkey, the US, the UK, & Germany

**Graphical abstract:** 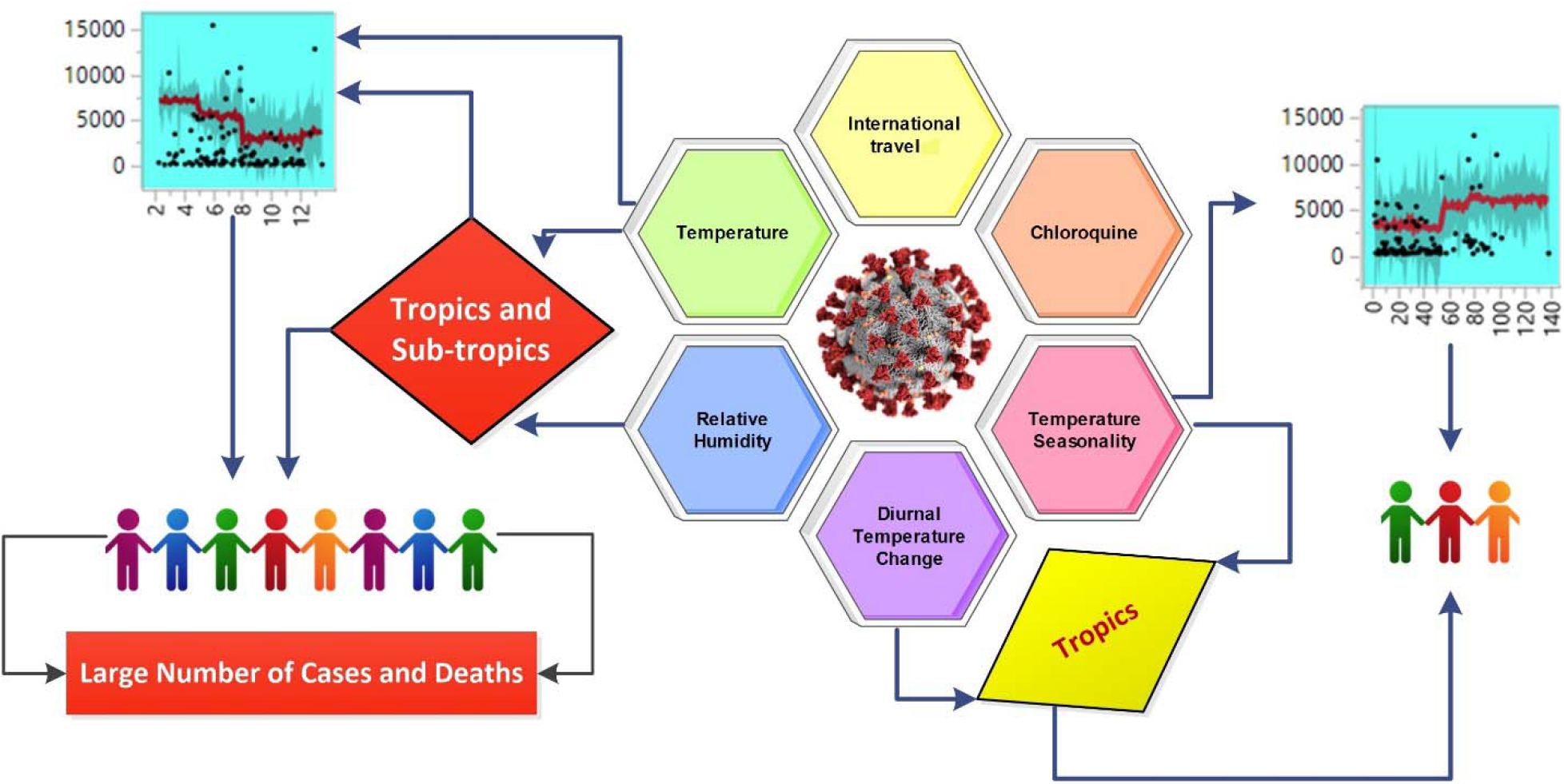

## 1. Introduction

The global surge of pandemic^1^ Severe Acute Respiratory Syndrome (SARS) coronavirus disease COVID-19) has been unprecedented in the 21^st^ century. The virus has spread rapidly across international borders^1^ through global travel from its primary infection epicenter in Wuhan^2^ (China) to new epicenters in Europe (Italy, Spain, France, Germany, the UK) and North America (the US and Canada). COVID-19 is highly contagious. The risk of human to human transmission is very high and mainly through close contact and respiratory droplets^2^,^3^ High fever, contagious cough, choking, severe pneumonia, and acute respiratory distress syndrome are the common symptoms.^3^ The case fatality rate (CFR) is estimated at 3.4%, while it varies by countries and population groups.^2^ The CFR of the current SARS-COV-2 is lower than its predecessor SARS-COV-1^3^, but its reproduction rate is much higher.

SARS-CoV-1 outbreak in 2003 infected more than 8000 individuals from 29 countries, and 774 died within a period of eight months, whereas, the COVID-19 has currently infected more than four million people across 212 countries with a death toll of close to 280,000 within four months.^4–6^ The very high infection susceptibility or high reproduction rate^4^ of this virus makes it particularly dangerous to older people, especially when the vaccinations and the drugs for treatment are not available.^7^

Historical evidence shows that meteorological conditions, e.g., temperature and relative humidity bring changes into the human activities that can influence more infections by increasing the reproduction rate of a virus.^8^ For instance, the higher air temperature may lead to an increase in the use of centrally air-conditioning systems, which host and spread the bacillus, causing Legionnaires’ Disease.^9^ Besides, the differential climatic conditions also lead to changes in the incidences of various infectious diseases such as malaria^10^, dengue^11^, influenza^12^, meningococcal meningitis^13^, cryptosporidiosis^14^, Rift Valley Fever^15^ Kyasanur Forest disease (KFD)^16^ and Lyme disease.^17,18^

Many studies suggest that the climatic conditions, e.g., humidity and temperature play key roles in spreading infectious diseases including SARS-COV-1, 2003.^19,20^ The daily incidence rate (DIR) of SARS-COV-1 was 18 times higher in low temperature than that higher temperature.^21^ Moreover, high circulation of influenza viral diseases has been found in the winter season in the temperate region of the southern and northern hemispheres.^22,23^The relative humidity is also a leading cause of occurrences of the influenza epidemic in the US and Vietnam.^24,25^

Few recent studies argued that meteorological factors, e.g., humidity and temperature could drive the pace of ongoing COVID-19 infections^26–28^ and local climatic conditions may drive COVID-19 growth rate.^29^Yet, the scientific community lacks evidence regarding the potential associations between climatic factors and COVID-19 cases at the global level. Most of the previous studies rely on the evidence from the regional levels of data and limited climatic variables. Luo et al. (2020)^30^ examined the relationship between province-level climatic variability and increase of COVID-19 reported cases and suggested that without extensive public health interventions, increase in temperature and humidity will not lead to a decline in COVID-19 cases. More importantly, Oliveiros et al. (2020)^28^ signify predictors percentage contribution in the rate of progressions of COVID-19 cases in which temperature and humidity only contribute to 18% and remaining 82% related to other factors, such as public health, population, infrastructure. Therefore, it is essential to determine the role of climatic factors (e.g., relative humidity and average temperature) behind the spread of COVID-19, to strengthen the knowledge base of COVID-19 research.

As the virus spread across the globe, the number of international travellers is the primary predictor of COVID-19 outbreak^31^ at national, regional, and local/city level. Due to high community transmission efficiency, the global cases are increasing day by day.^4^ However, there is a significant variation in the number of COVID-19 cases in terms of growth rate and timing around. Seoul (South Korea), Tokyo (Japan), and Bangkok (Thailand) appear to have been able to “flatten the curve”. At the same time, in several other countries (i.e., India and Brazil) in the tropical region, the number of COVID-19 cases are reported to be increasing significantly. Considering the above, ongoing COVID-19 pandemic situations and its increasing growth rate, more systematic research is essential, which accounts for climatic predictors, international travel, and chloroquine distribution information. In this background, the present study aims to identify the relationship between the efficiency of community transmission (spread) of the number of COVID-19 cases and climatic and bioclimatic factors as well as international travel information and public health concerns across severely affected cities distributed across tropics, sub-tropics and temperate climatic zones.

## 2. Methods

### 2.1 Selection of study sites

The international travelers were the primary cause of the spread of the COVID-19 to the global cities. The cities are more susceptible to the spread due to more substantial human mobility, service sector engagement, and tourist visitors as compared to the rural areas. Therefore, cities are the primary focus of the present study. Also, COVID-19 cases vary significantly from one country to another, and the month of the transmission is also different globally.

Therefore, to understand the pattern of the efficiency of region-wise community transmission, we collected the data for the countries where more than five cities were found to be significantly affected by COVID-19 cases with a higher increasing rate as of 21 April 2020. We have selected at least three cities with the most cases from each country across the world. In the case of the countries with the largest spillover, including the US, Spain, Italy, France, Germany, the UK, Turkey, Russia, Brazil, and India, the study selected ten cities for the analysis from each country. Further, out of ten cities, we have chosen five cities with the highest number of cases, whereas the remaining five cities were selected randomly to reduce the biases in the representation of a particular country. For remaining all the countries where cases are medium or less, we have taken one to three most affected cities based on the area of the countries. For smaller countries, one city, and for medium or larger countries, we have considered three cities as representatives for those countries.

A total of 230 cities were selected for the present study. To understand regional differentiation of COVID-19 cases, the cities were divided into tropical (0–23°26′11.9″ N/S), subtropical (23°26′11.9″ N/S- 40° N/S), and temperate (40° N/S – 60°N/S) zones based on latitudes. In the present study, 72, 63, and 93 cities were located in tropical, subtropical, and temperate regions, respectively. Two cities with polar climate were excluded from the study. The study used Boosted Regression Tree (BRT) model across climatic regions and larger spillover countries. In the following sub-section, details on variable selection and measurement, data collection procedure, and model specification are briefly described.

### 2.2 Descriptions and measurement of predictors

The present study collected and compiled the number of COVID-19 cases data at the city level from the WHO situation reports, health websites of different countries, and some data were also collected from the news bulletin, where all cases were regularly updated.

Air temperature and absolute humidity are two critical variables that may contribute to higher community transmission.^32^ In the context of COVID-19, the survival and transmission rates of viruses are mostly higher in the regions with low humidity and cold temperature.^29^ Hence, it was hypothesized that the higher the relative humidity and temperature, the lower the number of coronaviruses cases. Therefore, for the present analysis, the study used temperature and temperature-dependent bioclimatic variables (e.g., average diurnal temperature range, minimum temperature of the coldest month, average temperature of the coldest quarter, and temperature seasonality) and relative humidity as predictors. For each city, we extracted the average monthly temperature, and the average relative humidity data from the ECMWF ERA-5 reanalysis for January to April 2020.^33^ The month with a maximum number of reported cases was considered to tabulate temperature and relative humidity predictors data for the respective countries. The bioclimatic data of all selected countries were extracted from the worldclim historical dataset with a 1km resolution. To control for over-dispersion, we choose the maximum reported cases based on climate for the month, for example January for China and March for Italy.

Several studies showed that Chloroquine, a widely-used anti-malarial drug has a potentiality to reduce the vulnerability of COVID-19.^34–36^ Chloroquine is generally used for the prevention of malaria and is beneficial for treating rheumatoid arthritis in the anti-inflammatory patient. Chloroquine anti-viral and anti-inflammatory activities may be efficient in the treatment of COVID-19 patients.^34,37^ Malaria cases are calculated by the Annual Parasite Index (API) using both parasites, viz, Plasmodium vivax (Pv), and Plasmodium falciparum (Pf). However, in malaria-affected regions, Chloroquine is the main drug to alleviate symptoms, which can explain the reason behind fewer cases in African and South Asian countries. Hence it is assumed as one of the most important predictors to explain the number of COVID-19 cases worldwide. The API data were then extracted from the worldwide malaria cases reported in a study of Battle et al. (2015).^38^

It is also established that countries with a higher number of international travelers are more likely to have high potentiality to infect other people.^39^ As the data on international passengers are not available for 2019 and 2020, we used the 2018 data as a proxy to capture the traffic of international passengers in selected countries. The international passenger data were collected from the World Bank database.

### 2.3 Modelling approach

We analyzed the cases using a BRT model across the climatic regions and the countries with a large number of cases. BRT is an additive stochastic model that integrates regression trees by including an outcome to their predictors by recursive binary splits and combining multiple models to a single model, optimizing the predictive performance.^40^ The model can describe non-linear changes, accommodate missing data, and overcome the problems of outlier data.^41^ BRT models are found to be robust for a small number of data with missing data.^42^ BRT model can describe multiple interaction, partial dependency (non-monotonous and non-linear) of predictors, with sufficient flexibility and very high predictive accuracy. Therefore, the model is used to capture the non-linear relationship between the number of COVID-19 cases and selected variables in the present study.

To run the BRT model, we first evaluated the multicollinearity using Pearson correlation coefficient (r) and r ≥ 0.85 was selected as a cut-off threshold **(Figure 1; Table S1)** to remove the less important variables.^43^ The variables were cross-validated using the Variance Inflation Factor (VIF). We found that the VIF value is more than ten and insignificant for variables temperature of the coldest month, and an average temperature of the coldest quarter **(see Table S2)**, and hence these two variables were dropped from the analysis.^44^ COVID-19 cases were selected as outcome variable along with a set of six independent variables or predictors **(see Table 1):** average temperature, diurnal temperature change, temperature seasonality, relative humidity, number of travelers, and API.

**Table 1:** List of preliminary and final selected variable for the BRT model.

| Sl. no. | Predictors for the COVID-19 | Preliminary selected variables | Variable for the final model |
| --- | --- | --- | --- |
| 1. | Average temperature (°C) | * | √ |
| 2. | Monthly relative humidity (%) | * | √ |
| 3. | Diurnal temperature change (°C) | * | √ |
| 4. | Temperature Seasonality (%) | * | √ |
| 5. | Mean temperature of the coldest month (°C) | * | - |
| 6. | Mean temperature of the coldest Quarter (°C) | * | - |
| 7. | No of Passengers | * | √ |
| 8. | API values (per 1000 pops) | * | √ |

**Fig 1:**
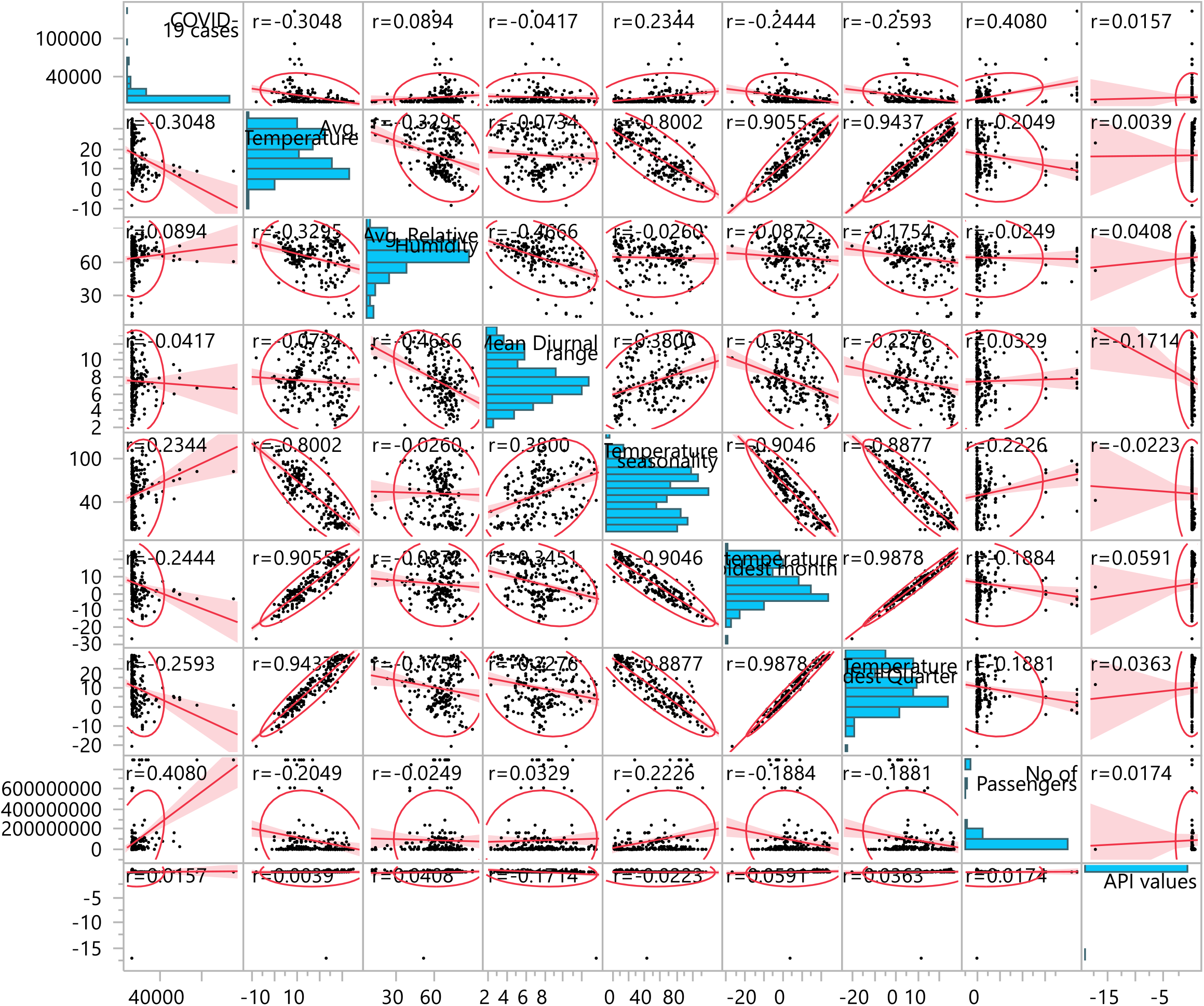
Scatterplot matrix showing the relationship between preliminary predictors and the number of COVID-19 cases. The corresponding correlation value (r) to identify insignificant predictors for the model are shown on the subplots.

#### 2.3.1 Boosted regression tree modelling

The motivation for boosting regression was to improving various weak learners by combining two powerful procedures: regression tree and boosting.^40,45^ The following gradient boosting model considers the forward stage-wise manner by adding the trained model from F, an approximation function of the response variable.

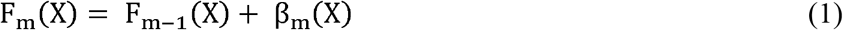

where β_m_ (X) is a weak learner of the basic functions. In the BRT model, β_m_ is the small regression tree and F_m_(X) is the sum of the small regression trees. For each boosting interaction, m number of new regression trees are added to the BRT model (m = 1, 2, 3…….M). The input x denotes the predictors from **Table 1** (finally selected variable), aims to estimate response Y_i.t+k_ from a training set, which entails the perfect β_m_ to satisfy F

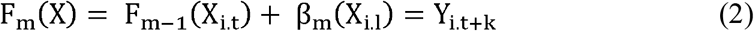

which is similar to

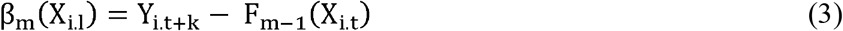

β_m_ in equation (3) is the current residuals r_m.i.t_ = Y_i.t+k_ – F_m-1_(X_i.t_) interaction with m to notice that current residuals have a negative slope of square error of loss functions,

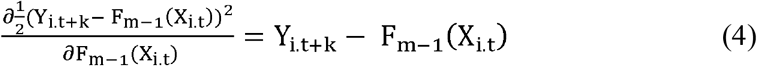

It indicates that β_m_ equalize the negative gradient of the squared loss function. Moreover, equation (4) proves that loss function is minimized in the gradient boosting algorithm. It also generalizes other loss functions by substituting the square error with different loss functions and their gradients. For more details of BRT, see Hastie et al. (2011), Scikit-learn (2015), Persson et al. (2017)^45–47^.

To avoid overfitting, a simple regularization strategy is to scale the contribution of each regression tree by a factor v.

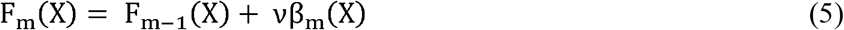

The parameter ν is the learning rate as it magnifies the length of the gradient descent procedure. strongly interacts with the number of boosting iterations M. In order to converge training error, smaller values of require more iterations and more basic functions. Several studies suggest that low values of favor better test error. For a more detailed discussion of the interaction between and M, see Ridgeway (2007).^48^

#### 2.3.2 Model Calibration

In this model, a 25% sample were used for training, and 75% sample distributed for testing. This method has been simulated 1,000 times to generate statistical inference by using ten times the loss function by cross-validation. In each BRT model, the subsampling procedure requires a parameter called the ‘bag fraction’ which was set at 0.75^49^, and at least 1,000 nodes/trees were used.^40^ In addition, a sensitivity analysis was conducted by setting a bag fraction of 0.5. All results presented in the following sections were calculated by averaging the predicted values of 50 bootstrap replicates. All analyses were conducted using DISMO package version Rv3.4.0. Moreover, the marginal association was assessed for all independent variables across climatic regions and the countries with major COVID-19 cases spillover. The relative contribution of response variables were also assessed, where a larger value indicated higher importance.^50^

#### 2.3.3 Model Validation

The model results were checked using the area in the Receiver Operating Characteristic (ROC) curve. Area under the ROC Curve (AUC) values differ between 0 and 1. The value of 0.5 suggests that the model results were less than random, and the value of 1.0 implies absolute discrimination.^43,51^

## 3. Results

### 3.1 Model validation and bag fraction analysis

The area under the curve in ROC (**Figure 2)** for the tested data was 0.9462, which confirms a high level of accuracy and forecasting ability of the model.^43^ A comparison between two bag fractions (0.5 and 0.75) was carried out in BRT models (Table S3). In general, only small variations within 2% were observed in relative contributions (RCs) of variables. The highest difference between RCs in temperature was about 1.87% in Russia **(Table S3)**.

**Fig. 2.**
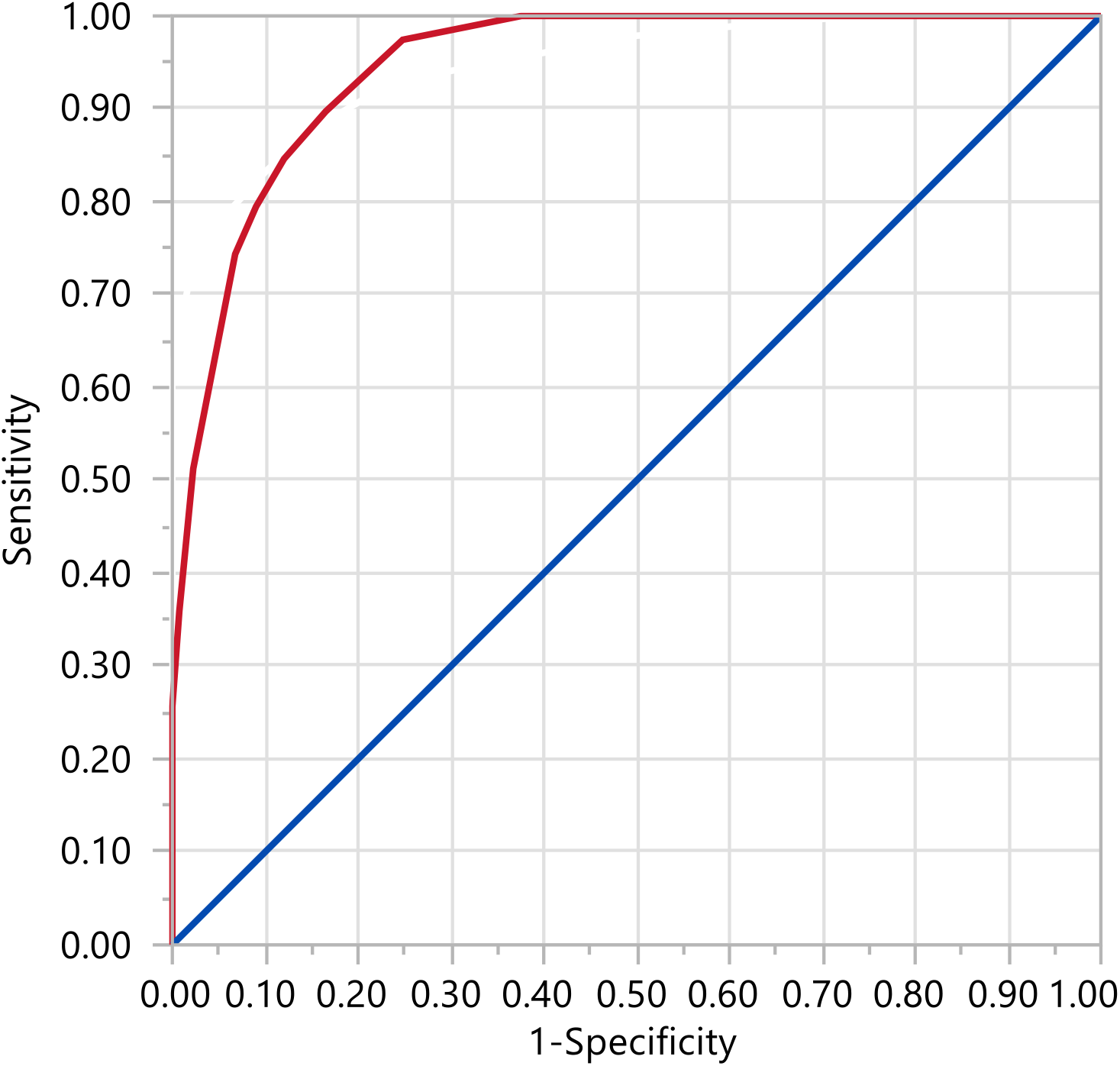
The ROC curve for the BRT model. The AUC value 0.9462 indicates that the forecasting ability for the model is very good, showing a high level of accuracy.

### 3.2 Descriptive statistics

As of 10 May 2020, a total of 4.18 million people were affected, and 0.283 million deaths were reported in the world.^52^ The virus has affected 210 territories and countries, wherein most of the cases were reported in developed countries. The climatic conditions may be relevant to the variation in the number of COVID-19 cases. To better understand the role of climatic predictors, **Table 2** shows the median, 10^th^ percentile and 90^th^ percentile of the average temperature, average relative humidity, diurnal temperature change, temperature seasonality in selected countries and regions across the globe. In the temperate zone, median average temperature, average relative humidity, diurnal temperature change, and temperature seasonality found to be 9°C, 67%, 7°C, and 70%, respectively, whereas 25°C, 65%, 7°C, and 27%, respectively, in the tropical zone **(Table 2)**. It indicates that there is a significant variation in temperature and temperature seasonality within these climatic regions. The number of COVID-19 cases are negatively associated with average temperature, diurnal temperature change, and relative humidity, and positively associated with temperature seasonality for all climatic region **(Figure 3 and 4)**. Also, the number of COVID-19 cases was positively associated with the number of international travelers and API **(Table 3)**.

**Table 2:** Meteorological quantiles (10th, 90th percentiles) of climatic, bioclimatic, and API values for COVID-19 cases in different climate zones and largest spillover countries in the World.

| Countries | Medians (10<sup>th</sup>, 90<sup>th</sup> percentiles) |  |  |  |  |
| --- | --- | --- | --- | --- | --- |
|  | Avg. temperature (°C) | Temperature seasonality (%) | Avg. relative humidity (%) | Mean diurnal range (°C) | API values (per 1000 pops) |
| The countries with the highest number of COVID-19 cases |  |  |  |  |  |
| USA | 10 (6,17) | 84 (47,93) | 65 (60,71) | 8 (7,11) | - |
| Spain | 12 (10,15) | 57 (48,66) | 70 (63,77) | 9 (6,10) | - |
| Italy | 11 (9,13) | 70 (63,72) | 63 (62,71) | 8 (6,9) | - |
| Germany | 8 (7,9) | 58 (57,66) | 66 (64,69) | 7 (7,7) | - |
| UK | 7 (6,9) | 44 (41,45) | 73 (67,76) | 6 (5,6) | - |
| Russia | 2 (0,4) | 111 (102,115) | 82 (75,89) | 8 (7,8) | - |
| Turkey | 11 (9,14) | 68 (67,84) | 69 (63,71) | 8 (5,11) | - |
| France | 10 (8,12) | 59 (54,64) | 68 (66,70) | 7 (5,8) | - |
| Brazil | 27 (25,28) | 22 (20,23) | 73 (67,77) | 8 (7,9) | 0.12 (0.06,0.18) |
| India | 28 (27,30) | 22 (20,55) | 59 (43,72) | 8 (5,10) | 0.005 (0.001, 0.009) |
| Climatic zones |  |  |  |  |  |
| Tropical | 25 (10,31) | 27 (8,80) | 65 (41,78) | 7 (4,11) | - |
| Sub-tropical | 17 (10,25) | 58 (33,96) | 64 (41,72) | 8 (5,12) | - |
| Temperate | 9 (4,17) | 70 (46,97) | 67 (57,77) | 7 (5,11) | - |

**Fig 3:**
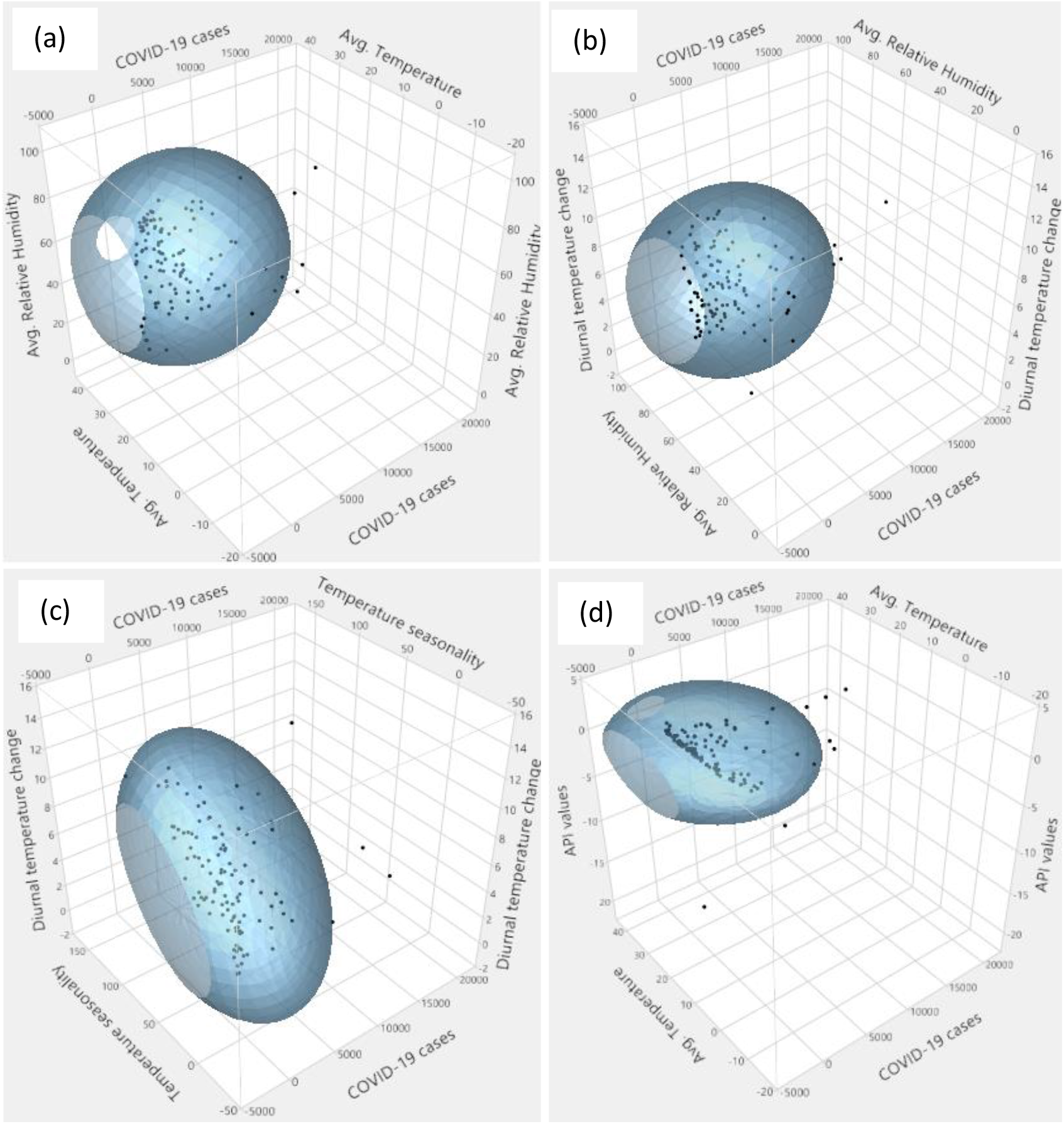
Relationship between selected climatic variables, number of international travelers and number of COVID-19 cases in the tropical region. Fig (a) COVID-19, relative humidity, and temperature; (b) COVID-19, diurnal range of temperature, and relative humidity; (c) COVID-19, temperature seasonality, and diurnal temperature change; (d) COVID-19, average temperature and API values.

**Fig 4:**
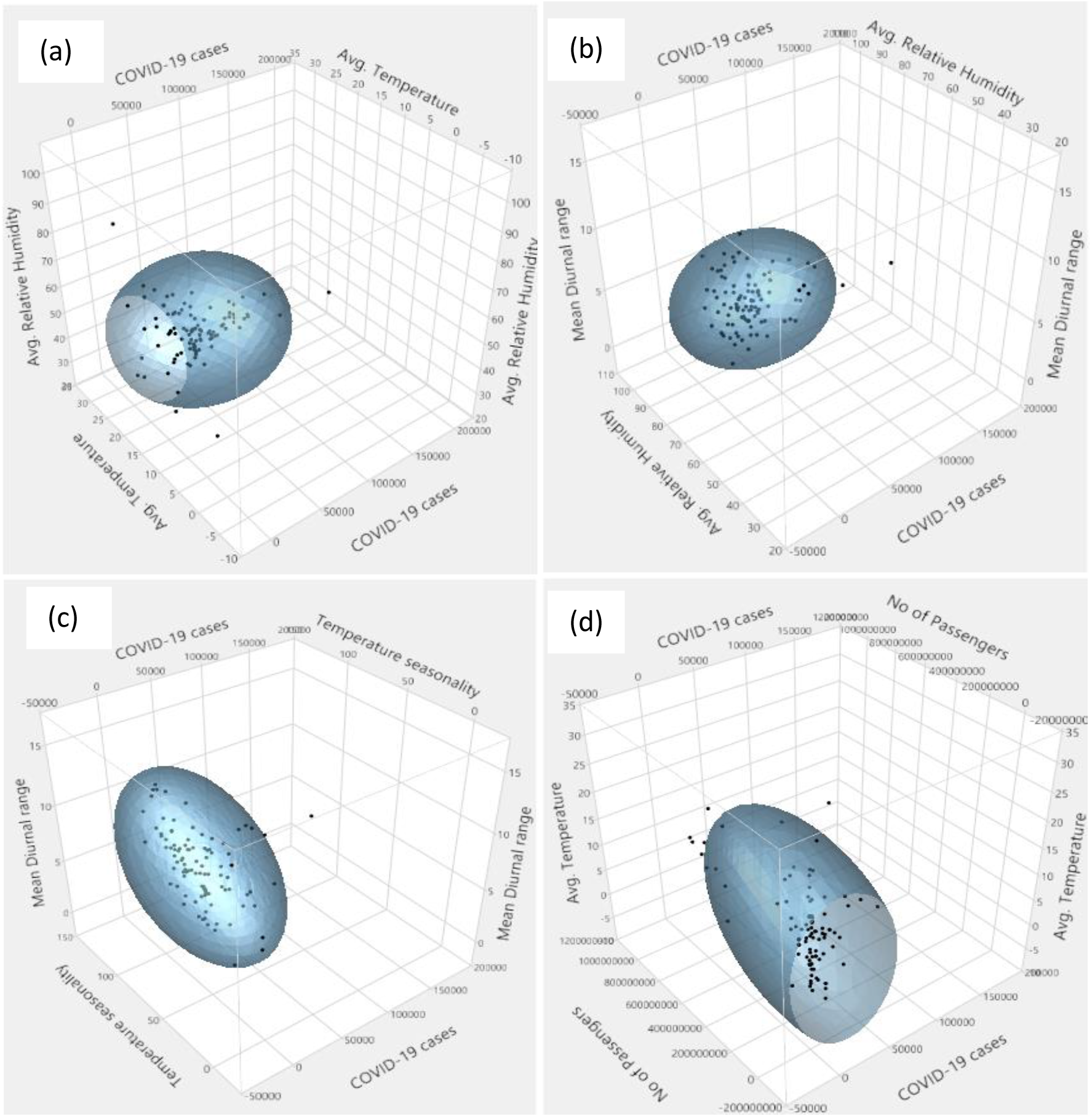
Relationship between selected climatic variables, the number of international travelers, and the number of COVID-19 cases in the temperate region, and a similar relationship has been observed for the sub-tropical region. Fig (a) CoVID-19, relative humidity, and temperature; (b) COVID-19, diurnal range of temperature, and relative humidity; (c) COVID-19, temperature seasonality, and diurnal temperature change; (d) COVID-19, average temperature and API values.

### 3.3. Relative effects of predictors

**Table 3** presents the association between COVID-19 and climatic parameters, number of international passengers and API based on aggregate global model. We excluded the information about the number of international travelers for country-level analysis due to single data for the country level. Although it is a primary source of infection, it has no role in community transmission within a country. For the variable of API, we considered only those regions which have malaria endemicity.

**Table 3:**
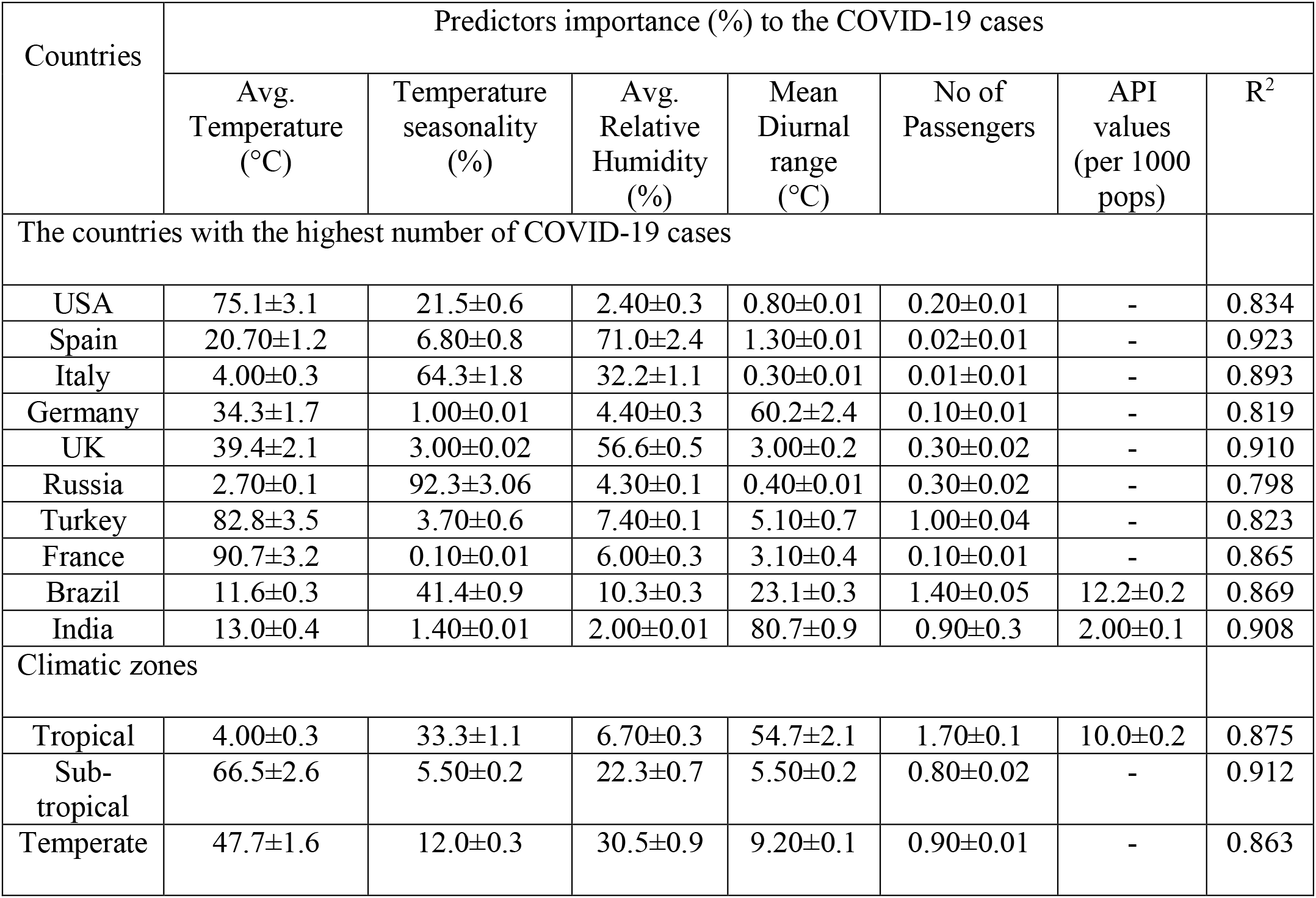
Relative importance of predictors (climatic, bioclimatic, travel passenger, and API variables) in percent (±SD) and goodness of fit of the model.

| Countries | Predictors importance (%) to the COVID-19 cases |  |  |  |  |  |  |
| --- | --- | --- | --- | --- | --- | --- | --- |
|  | Avg. Temperature (°C) | Temperature seasonality (%) | Avg. Relative Humidity (%) | Mean Diurnal range (°C) | No of Passengers | API values (per 1000 pops) | R<sup>2</sup> |
| The countries with the highest number of COVID-19 cases |  |  |  |  |  |  |  |
| USA | 75.1±3.1 | 21.5±0.6 | 2.40±0.3 | 0.80±0.01 | 0.20±0.01 | - | 0.834 |
| Spain | 20.70±1.2 | 6.80±0.8 | 71.0±2.4 | 1.30±0.01 | 0.02±0.01 | - | 0.923 |
| Italy | 4.00±0.3 | 64.3±1.8 | 32.2±1.1 | 0.30±0.01 | 0.01±0.01 | - | 0.893 |
| Germany | 34.3±1.7 | 1.00±0.01 | 4.40±0.3 | 60.2±2.4 | 0.10±0.01 | - | 0.819 |
| UK | 39.4±2.1 | 3.00±0.02 | 56.6±0.5 | 3.00±0.2 | 0.30±0.02 | - | 0.910 |
| Russia | 2.70±0.1 | 92.3±3.06 | 4.30±0.1 | 0.40±0.01 | 0.30±0.02 | - | 0.798 |
| Turkey | 82.8±3.5 | 3.70±0.6 | 7.40±0.1 | 5.10±0.7 | 1.00±0.04 | - | 0.823 |
| France | 90.7±3.2 | 0.10±0.01 | 6.00±0.3 | 3.10±0.4 | 0.10±0.01 | - | 0.865 |
| Brazil | 11.6±0.3 | 41.4±0.9 | 10.3±0.3 | 23.1±0.3 | 1.40±0.05 | 12.2±0.2 | 0.869 |
| India | 13.0±0.4 | 1.40±0.01 | 2.00±0.01 | 80.7±0.9 | 0.90±0.3 | 2.00±0.1 | 0.908 |
| Climatic zones |  |  |  |  |  |  |  |
| Tropical | 4.00±0.3 | 33.3±1.1 | 6.70±0.3 | 54.7±2.1 | 1.70±0.1 | 10.0±0.2 | 0.875 |
| Sub-tropical | 66.5±2.6 | 5.50±0.2 | 22.3±0.7 | 5.50±0.2 | 0.80±0.02 | - | 0.912 |
| Temperate | 47.7±1.6 | 12.0±0.3 | 30.5±0.9 | 9.20±0.1 | 0.90±0.01 | - | 0.863 |

**Table 3** represents the region and country wise association between climatic parameters and the number of COVID-19 cases. The results show that average temperature (47.70%) and average relative humidity (30.50%) were the major contributors in explaining the differentials of COVID-19 transmission in the temperate zone. At the same time, the mean diurnal range (54.70%) and temperature seasonality (33.30%) were the most significant determinants of this viral community transmission in the tropical zone. In the temperate zone, the role of average temperature (66.5%) and relative humidity (22.3%) were the highest among the selected all predictors.

The results show that in temperate countries, the average temperature was a major contributor to the number of cases in France (90.70%), Turkey (82.80%), the US (75.10%), the UK (39.40%), Germany (34.30%). Similarly, the average relative humidity contributed more in Spain (71.0%), the UK (56.60%), and Italy (32.20%), and favorable relative humidity for the spread was found in the range of 60 to 70 % in countries from the temperate zone. The Russian cases were mostly affected by the temperature seasonality contributing 92.30% to the spread. The mean diurnal temperature range was contributing about 60.20% of the cases in Germany **(Table 3)**.

The cities located in the tropical zone, like cities form India and Brazil, were mostly influenced by the diurnal temperature range. In India, 80.70% of the cases were explained by the mean diurnal temperature, followed by the average temperature (13%) and temperature seasonality (1.40%). The maximum number of cases in India was explained in the range of temperature seasonality 22% to 38%. The community transmission in Brazil was mostly influenced by temperature seasonality (41.40%), followed by the mean diurnal range of temperature (23.10%), API (12.20%) average temperature (11.60%), and relative humidity (10.30%) **(Table 3)**.

### 3.4 COVID-19 response to the predictors in different climatic regions

The association between climatic indicators and COVID-19 risks is illustrated in **Figure 5**. A non-linear relationship is observed between average temperature and COVID-19 cases in the temperate and sub-tropical zones. The results show that average temperature was negatively associated with COVID-19 transmission risks, which tend to reduce significantly when the average temperature varied from 5°C to 12°C in the sub-tropical zone and 5°C to 11°C in the temperate zone. With increasing average temperature, community transmission is reduced significantly. The response of the number of COVID-19 cases was slightly positive and associated with relative humidity, although it was a less influencing factor in the temperate and sub-tropical zones. After the threshold of about 60% relative humidity in these two regions, the probability of disease transmission increased **(Figure 5)**.

**Fig. 5:**
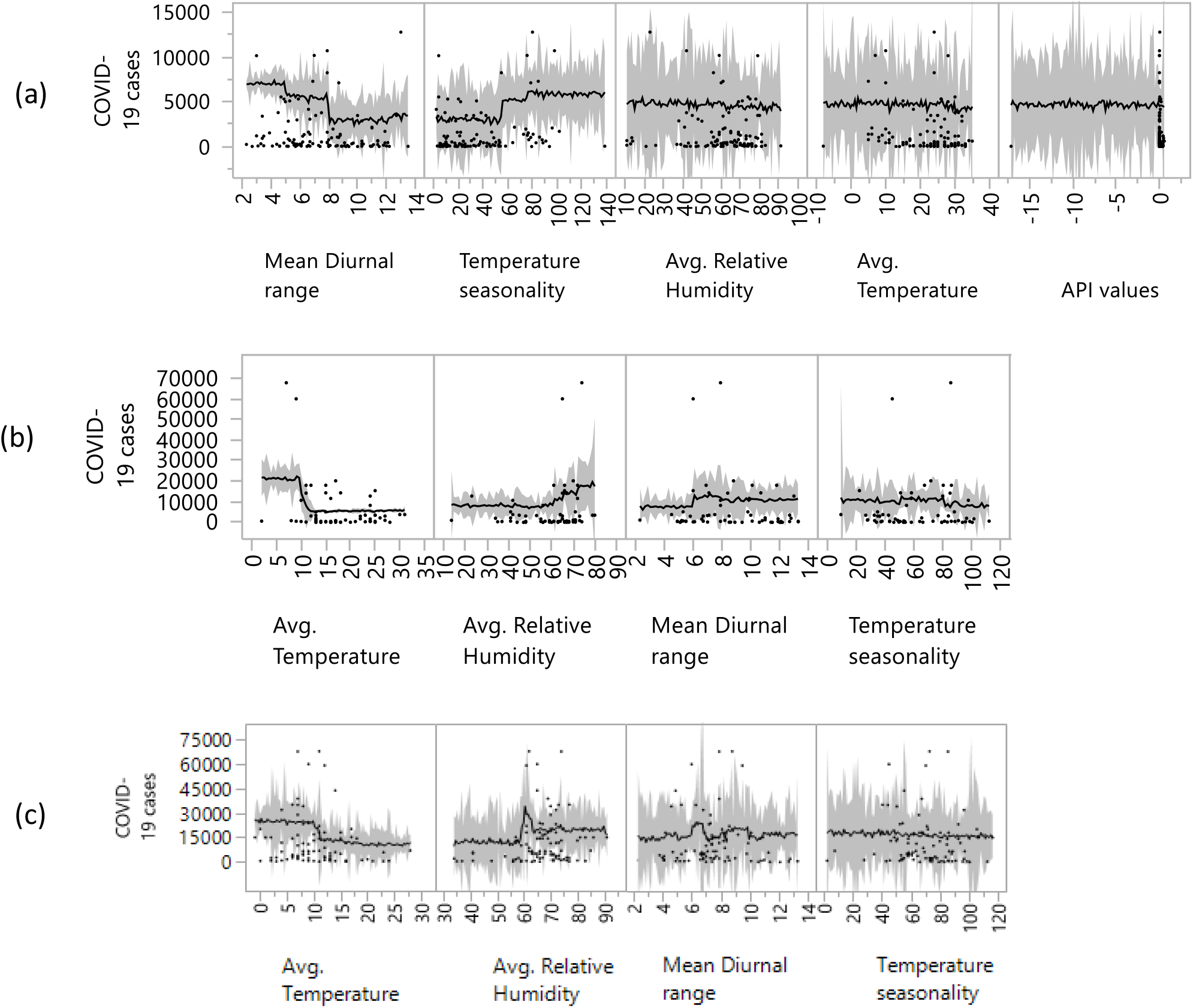
Marginal dependence graphs for the five most influential predictors in the model for COVID-19 in tropical (a), sub-tropical (b), and temperate (c) regions. For explanation of predictors and their units, see Table 3. Y*-*axes are showing number COVID-19 cases, and X-axes represent predictors. The predictor, the number of international travelers, was omitted as this is a primary source of infection, but it has no role in community transmission. The API values considered only for the tropical region, where most of the countries of this region are malaria-prone. The shaded line shows a 95% confidence interval from the mean.

On the contrary, these two meteorological parameters did not have a significant association with the disease transmission in the tropical region. The significant community transmission occurred with the changes in mean diurnal temperature, which was ranged from 4 to 8°C. After this, there was a significant decline in the number of COVID-19 cases in community transmission, which had little variations with average temperature. The temperature seasonality was also a significant variable showing positive association for the community transmission in the tropical countries. Besides these factors, API had a very poor association with the COVID-19 transmission in the tropical regions.

### 3.5 COVID-19 response to the predictors in different countries

**Figure 6** represents the country-wise association between the climatic predictors and the COVID-19 cases. The results show that in France, Turkey, the US, the UK, Germany, the number of COVID-19 cases were non-linearly but highly associated with average temperature. Maximum cases were found during the temperature range of 5 to 10°C, and after the temperature increased beyond 10°C infected cases declined. Similarly, the average relative humidity was a contributing factor in Spain, the UK, and Italy, and favorable relative humidity for the disease transmission was found to be 60 to 70% in temperate countries. Most interestingly in the case of Turkey, it was found that the cases were increasing after crossing the 73% threshold of relative humidity. The temperature seasonality mostly influenced the Russian cases. About 92% of the cases in Russia were influenced by temperature seasonality, followed by Italy (64.3%), and the US (21.5%). It concludes that more than 70% variation of temperature (temperature seasonality) may cause a significant increase in COVID-19 community transmission. But with the 80% of temperature seasonality, there was a declining trend for the US cases, whereas Russian cases declined after the value reached 110%. This might be because the location and extension of Russia is northwards than the US where extreme seasonality was found. Another important variable, mean diurnal range of temperature contributes more (about 60%) to the community transmission in Germany **(Figure 6)**.

**Fig. 6:**
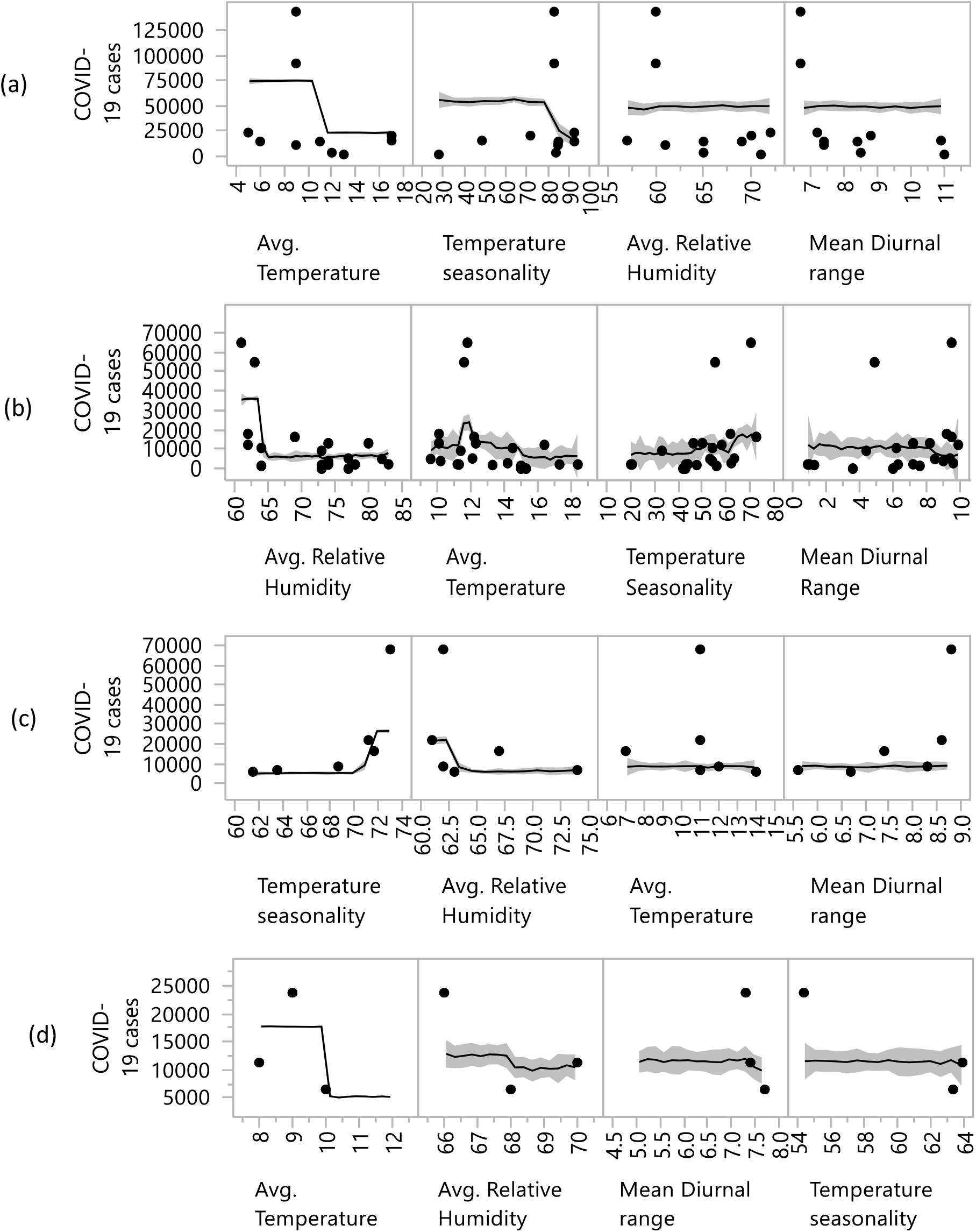
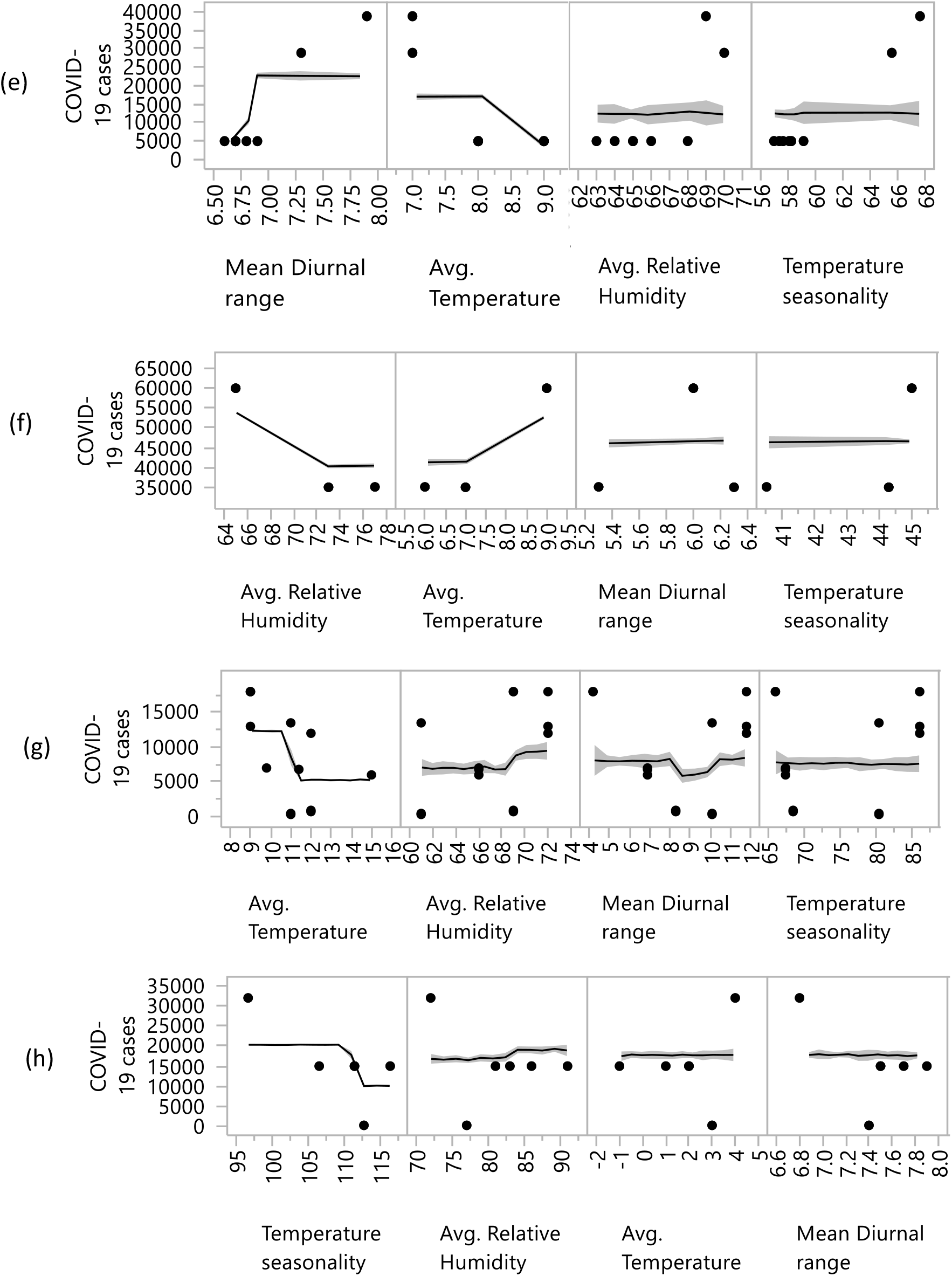
Marginal dependence graphs for the four most influential predictors in the model for COVID-19 disease in the USA (a), Spain (b), Italy (c), France (d), Germany (e), UK (f), Turkey (g), Russia (h). For explanation of predictors and their units, see Table 3. Y-axes are showing the number of COVID-19 cases, and X-axes represent predictors. The predictor, the number of international travelers, was omitted as it is a primary source of infection, but it has no role in community transmission. The API was omitted for fewer malaria cases in the temperate region. The shaded line shows a 95% confidence interval from the mean.

It was found that the COVID-19 community transmission in the tropical zone was not strongly associated with the temperature. Maximum cases are explained by 30–40% of seasonal variation in temperature, and after 40% seasonal variation in temperature, the number of cases may decline sharply. The cases in India were mostly associated with the diurnal range of temperature (80.7%). The cases in Brazil were mostly influenced by the temperature seasonality (41.4%). In Brazil, the maximum number of cases were found in the range 5° to 8°C of the mean diurnal temperature. The API values were influenced slight positively, although the rate was very low. In Brazil, the cases were sharply increasing with an increase of average temperature. The results also showed that the average temperature ranging from 25 to 30°C was the most influential factor behind the number of cases in these tropical countries **(Figure 7)**.

**Fig. 7:**
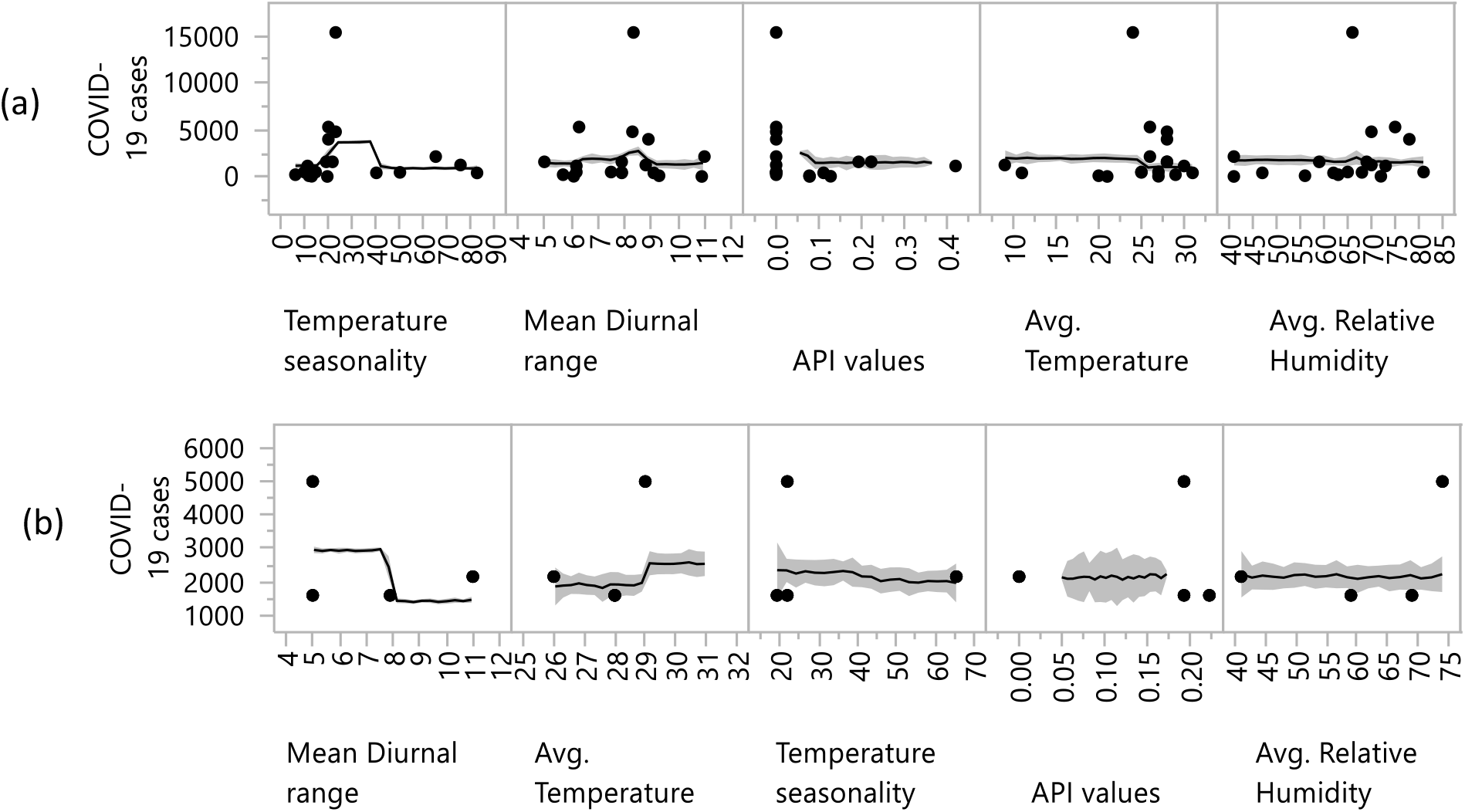
Marginal dependence graphs for the five most influential predictors in the model for COVID-19 in India (a), Brazil (b). For explanation of predictors and their units, see Table 3. Y-axes are showing COVID-19 cases, and X-axes represent predictors. The predictor, the number of international travelers, was omitted, as it is a primary source of infection, but has no role in community transmission. The value of API was considered in the analysis as these countries are malaria-prone. Shaded line shows a 95% confidence interval from the mean.

## 4. Discussion

Many studies have tried to establish the relationship between meteorological parameters and transmission of influenza epidemic.^12^ In recent times, several efforts have also been made to evaluate the association between climatic predictors and COVID-19 transmission.^26–29, 53^ Existing studies mainly focused on regional perspectives of COVID-19 transmission and its association with climatic conditions. However, studies at the macro level are limited, in particular, the studies which accounts for different climatic regions. Besides, the influence of climatic predictors, including travel information and API on the number of COVID-19 cases and community transmission, has limitedly established. An empirical analysis of the influence of climatic, bioclimatic, and factors like international travel information and API on COVID-19 community transmission using city-level data across three climatic regions in countries with the most number of COVID-19 cases is expected to improve the understanding of the spread of the disease.

The results of country-level analysis showed that in Indonesia, the only average temperature is linked with the COVID-19 transmission, while humidity, maximum or minimum temperatures are not correlated.^53^ In this background, the present study used climatic, bioclimatic and international travel information and API data for 72 cities from the tropical, 72 cities from the sub-tropical and 93 cities from the temperate zones. The study found that increasing temperature and decreasing average relative humidity were associated with the slowdown the community transmission of COVID-19. At the same time, Wang et al. (2020a)^26^ revealed that higher average temperature and higher relative humidity considerably decrease the COVID-19 transmission. About 1°C rise in average temperature is related to a reduction of reproduction rate of transmission by 0.0225 in China^26^ and a 1% rise in relative humidity lowers the reproduction rate by 0.0158. Another study by Bu et al. (2000) concludes that in China, average temperature ranges between 13°C and 19°C and average relative humidity ranges between 50% – 80% constitute an appropriate condition for the community transmission of this virus.^54^

A study from China showed that the cases of COVID-19 were highest within the 10°C while it is considerably low more than 10°C temperature.^27^ The present study found a linear relationship between the transmission of COVID-19 and temperature in the temperate region, while there was no significant association between these two in the tropical region. As China is from a temperate region, with an increase in temperature, the number of COVID-19 cases also increased in the country. It might therefore appear that COVID-19 needs a 4°C of minimum level of temperature for smooth transmission. Also, in the temperate and subtropical regions, COVID-19 transmission was lower when the temperature remains below 10°C. Possibly, in these regions, the unfavorable temperature keeps people inside their homes, and “social distancing” was maintained. Therefore, the temperature might have played a significant role in the dispersion of the virus in the temperate and subtropical regions.^55^ While the average temperature was not associated with COVID-19 transmission in the tropical region, the temperature seasonality and mean diurnal temperature become important for the transmission in the region. Since various parameters of temperature were associated differently with the outbreak in different climatic regions such as the temperate and tropical zones, it may also vary over regional/country levels due to changes in geographical and ecological settings. Thus, the regional level analysis of heterogeneous climatic associations with the transmission is equally necessary along the global assessments.

The present study found that the role of average relative humidity on COVID-19 transmission was weaker and inconsistent compared to the temperature. COVID-19 community transmission in temperate zone were generally suitable for growth in the number of cases in the conditions of high relative humidity but not exceedingly wet environments (>90%). Moreover, in the tropical zone, high relative humidity is also linked with the transmission rate of COVID-19 cases but not strongly associated, as in the temperate zone. The results of the present study are consistent with the previous studies, showing the inconsistent effects of relative humidity on COVID-19 cases in the regional case of China.^27^ The study also found a similar relationship for Hemorrhagic fever with renal syndrome (HFRS) in China, which was positively associated with cold days in China.^56^ The relationship between relative humidity and COVID-19 cases can be complicated in a country-level analysis as wet condition may block the viral replication.^12,55^ Deyle et al., (2016), signified that the effects of relative humidity on influenza disease depends on the temperature.^57^ This could explicate our findings that the impact of humidity on COVID-19 transmission could be stronger in the temperate zone and weaker in the tropical zone as a procession of seasonal temperature change.

More detailed country-specific findings revealed similar results to those of the regional level, albeit with slight variations. In most of the temperate countries such as France, the USA, Turkey, the UK, and Germany, the cities having an average temperature in the range 5–10°C have a higher level of COVID-19 transmission rate than their counterparts **(Figure 5)**. Besides, other climatic parameters like average relative humidity played an important role in some of the countries such as Italy, Spain, the UK, and Russia. In humid region with a favourable humidity 60–70%, if infected people sneezes and coughs, the released tiny droplets into the surrounding environment, and it travels further into the air. The droplets in the air may not evaporate soon, and more likely to infect a new people.^58^ In summary, temperature and humidity can be used for predicting the COVID-19 transmission in these countries.

Besides the climatic factors, our results showed that chloroquine distribution affects the COVID-19 transmission, particularly in tropical countries such as Brazil and India. A non-randomized clinical trial shows that anti-malarial drugs such as Hydroxychloroquine and azithromycin weaken the symptoms of COVID-19.^35^ The API values slightly positively influenced the number of COVID-19 cases in these countries, and the rate of influence was very low because cities are less prone to malaria globally.^38^ Hence, in the areas where the prevalence of malaria was relatively high, the percentage of infected people might be less in the form of asymptomatic or mild symptomatic as compared to the areas where the prevalence of malaria is low (e.g., South Asian and African countries). The development of immunity against malaria perhaps lessens the probability of showing symptoms among the people in malaria-affected regions.^35^

Other strains of coronavirus such as HCoV-HKU1, HCoV-229E, HCoV-OC43, and HCoV-NL63 generally show symptoms like the common cold. The COVID-19 seemed to have a strong seasonality effect from December to April, although data for other months are not available for the comparison. The transmission of the virus lessens during the summer season.^59^ In the coming months, in general, the temperature will be increasing in the countries from the northern hemisphere. At the same time, the temperature will be decreasing in the countries of the southern hemisphere. Hence, the findings from this study would have important implications in formulating strategies to deal with COVID-19-related in the near future. It should be noted that the present study does not predict the climatic parameter-based months with higher risk for the cities of different climatic zones. Future studies may emphasize on predicting the monthly climatic conditions and associated transmission risk of COVID-19 across the countries and regions. The present study does not include other factors, such as the human physiological response of a community to the virus and social and economic determinants of viral transmission due to data limitations.

## 5. Conclusions

The present study used city level climatic, bioclimatic, travel, and chloroquine distribution data to identify the relationship between the climatic region-wide and country-wide variations and the number of COVID-19 cases by the marginal effects of predictors. The study concludes that climatic and bioclimatic predictors across three climatic zones significantly affects the spread of the number of COVID-19 cases. The findings of the present study are expected to improve the understanding of the relationships between the climatic variables and the number of COVID-19 cases. It underlines the importance of meteorology-based early warning systems to facilitate timely response to COVID-19 community transmission. The finding from the present study are expected to add to the ongoing debates on the influence of climatic factors on the spread of COVID-19 cases and could help researchers and policymakers to make appropriate decisions for preventing the spread.

## Data Availability

The data are available on request

## Footnotes

1 On 30 January 2020, the COVID-19 announced as an Public Health Emergency of International Concern and on 11^th^ march, 2020 declared it as a pandemic.

2 Originated from the Wuhan fish market in December, 2019.

3 Severe acute respiratory syndrome coronavirus (SARS-CoV), 2002–04 epidemic. More than 8,000 people were infected from 29 countries, and 774 died worldwide

## Supplementary tables

**Table S1:** The Pearson correlation test results for COVID-19 cases and selected variables (5% significance level). The cut-off threshold is 0.85.

| Variables | COVID-19 cases | Avg. temperature | Avg. relative humidity | Mean diurnal range | Temperature seasonality | Mean temperature of the coldest month | Mean temperature of the coldest quarter | Number of passengers | API values |
| --- | --- | --- | --- | --- | --- | --- | --- | --- | --- |
| COVID-19 cases | 1.0000 | -0.3048 | 0.0894 | -0.0417 | 0.2344 | -0.2444 | -0.2593 | 0.4080 | 0.0157 |
| Avg. temperature | -0.3048 | 1.0000 | -0.3295 | -0.0734 | -0.8002 | 0.9055 | 0.9437 | -0.2049 | 0.0039 |
| Avg. relative humidity | 0.0894 | -0.3295 | 1.0000 | -0.4666 | -0.0260 | -0.0872 | -0.1754 | -0.0249 | 0.0408 |
| Mean diurnal range | -0.0417 | -0.0734 | -0.4666 | 1.0000 | 0.3800 | -0.3451 | -0.2276 | 0.0329 | -0.1714 |
| Temperature seasonality | 0.2344 | -0.8002 | -0.0260 | 0.3800 | 1.0000 | -0.9046 | -0.8877 | 0.2226 | -0.0223 |
| Mean temperature of coldest month | -0.2444 | 0.9055 | -0.0872 | -0.3451 | -0.9046 | 1.0000 | 0.9878 | -0.1884 | 0.0591 |
| Mean temperature of coldest quarter | -0.2593 | 0.9437 | -0.1754 | -0.2276 | -0.8877 | 0.9878 | 1.0000 | -0.1881 | 0.0363 |
| Number of passengers | 0.4080 | -0.2049 | -0.0249 | 0.0329 | 0.2226 | -0.1884 | -0.1881 | 1.0000 | 0.0174 |
| API values | 0.0157 | 0.0039 | 0.0408 | -0.1714 | -0.0223 | 0.0591 | 0.0363 | 0.0174 | 1.0000 |

**Table S2:**
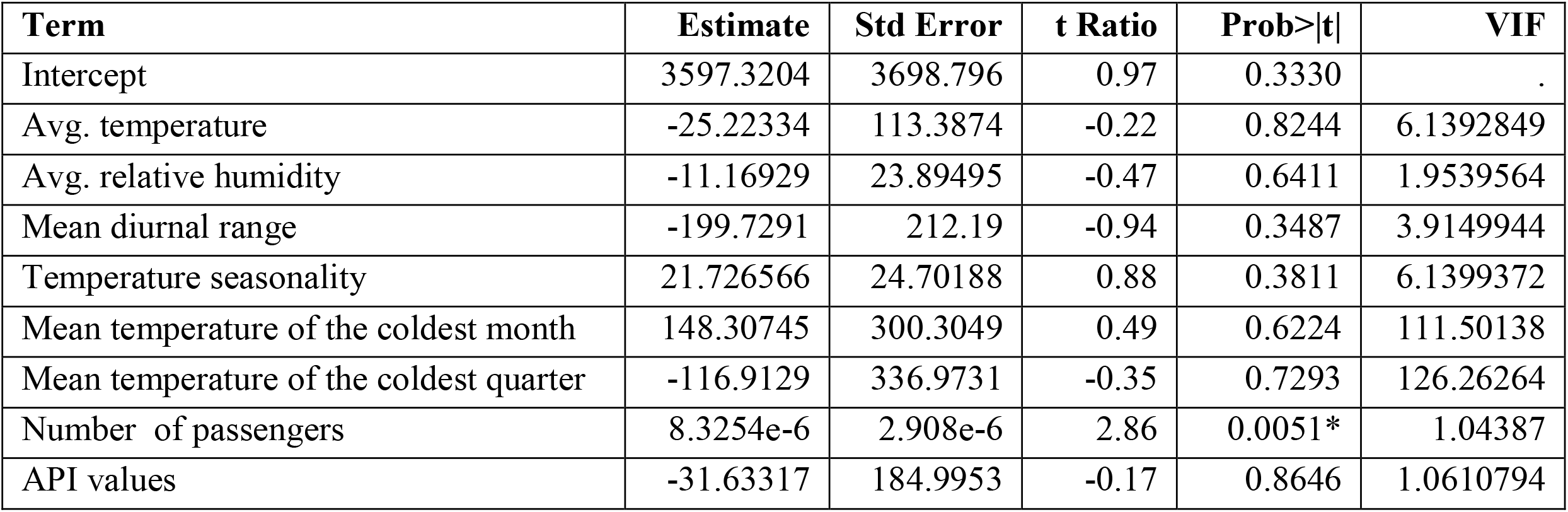
Showing Variance Inflation factor (VIF) value to check significant variable from the primarily selected variables

| **Term** | **Estimate** | **Std Error** | **t Ratio** | **Prob>\|t\|** | **VIF** |
| --- | --- | --- | --- | --- | --- |
| Intercept | 3597.3204 | 3698.796 | 0.97 | 0.3330 | . |
| Avg. temperature | -25.22334 | 113.3874 | -0.22 | 0.8244 | 6.1392849 |
| Avg. relative humidity | -11.16929 | 23.89495 | -0.47 | 0.6411 | 1.9539564 |
| Mean diurnal range | -199.7291 | 212.19 | -0.94 | 0.3487 | 3.9149944 |
| Temperature seasonality | 21.726566 | 24.70188 | 0.88 | 0.3811 | 6.1399372 |
| Mean temperature of the coldest month | 148.30745 | 300.3049 | 0.49 | 0.6224 | 111.50138 |
| Mean temperature of the coldest quarter | -116.9129 | 336.9731 | -0.35 | 0.7293 | 126.26264 |
| Number of passengers | 8.3254e-6 | 2.908e-6 | 2.86 | 0.0051* | 1.04387 |
| API values | -31.63317 | 184.9953 | -0.17 | 0.8646 | 1.0610794 |

**Table 3.** Model settings for the bag fraction (0.50 and 0.75) comparison for the relative importance of the selected predictors settings in the BRT models.

| Countries & climatic regions | Predictors importance (%) to the COVID-19 cases |  |  |  |  |  |  |
| --- | --- | --- | --- | --- | --- | --- | --- |
|  | Bag fraction | Avg. temperature (°C) | Temperature seasonality (%) | Avg. relative humidity (%) | Mean diurnal range (°C) | Number of Passengers | API values (per 1000 population) |
| USA | 0.75 | 75.1 | 21.5 | 2.4 | 0.8 | 0.2 | - |
|  | 0.50 | 74.4 | 20.9 | 2.3 | 0.7 | 0.2 |  |
| Spain | 0.75 | 0.70 | 0.21 | 99.0 | 0.07 | 0.02 | - |
|  | 0.50 | 0.60 | 0.22 | 98.2 | 0.06 | 0.01 |  |
| Italy | 0.75 | 4.00 | 64.3 | 32.2 | 0.3 | 0.01 | - |
|  | 0.50 | 3.80 | 63.1 | 31.1 | 0.2 | 0.01 |  |
| Germany | 0.75 | 34.3 | 1.0 | 4.4 | 60.2 | 0.1 | - |
|  | 0.50 | 33.9 | 0.8 | 4.3 | 59.6 | 0.07 |  |
| UK | 0.75 | 39.4 | 3.0 | 56.6 | 3.0 | 0.3 | - |
|  | 0.50 | 38.7 | 3.2 | 56.2 | 2.9 | 0.23 |  |
| Russia | 0.75 | 2.70 | 92.30 | 4.3 | 0.4 | 0.3 | - |
|  | 0.50 | 2.50 | 90.43 | 4.1 | 0.3 | 0.33 |  |
| Turkey | 0.75 | 82.8 | 3.7 | 7.4 | 5.1 | 1.0 | - |
|  | 0.50 | 82.6 | 3.8 | 7.2 | 4.9 | 0.8 |  |
| France | 0.75 | 90.7 | 0.1 | 6.0 | 3.1 | 0.1 | - |
|  | 0.50 | 90.5 | 0.1 | 6.3 | 2.9 | 0.8 |  |
| Brazil | 0.75 | 11.6 | 41.4 | 10.3 | 23.1 | 1.4 | 12.2 |
|  | 0.50 | 11.5 | 41.3 | 9.8 | 23.0 | 1.3 | 12.1 |
| India | 0.75 | 13.0 | 1.4 | 2.0 | 80.7 | 0.9 | 2.0 |
|  | 0.50 | 12.6 | 1.3 | 1.7 | 79.4 | 0.6 | 1.8 |
| Tropical | 0.75 | 4.00 | 33.3 | 6.7 | 54.7 | 1.7 | 10.0 |
|  | 0.50 | 4.10 | 33.4 | 5.8 | 53.4 | 1.3 | 10.2 |
| Sub-tropical | 0.75 | 66.5 | 5.5 | 22.3 | 5.5 | 0.8 | - |
|  | 0.50 | 66.1 | 5.6 | 21.3 | 5.3 | 0.6 |  |
| Temperate | 0.75 | 47.7 | 12.0 | 30.5 | 9.2 | 0.9 | - |
|  | 0.50 | 47.1 | 12.3 | 31.3 | 9.3 | 0.6 |  |

